# Multimodal Speech and Text Models to Detect Suicidal Risks in Adolescents

**DOI:** 10.1101/2025.07.31.25332367

**Authors:** Jiacheng Jin, Yuanfei Liu, Ying Dai

**Affiliations:** Department of Electrical Engineering, Zhejiang University, China; Department of Psychiatry, University of Cambridge, UK; Department of Nursing, Guangzhou Women and Children’s Medical Center, Guangzhou Medical University, China

**Keywords:** Adolescent, Classification, Computational paralinguistics, Speech recognition, Suicidal risks

## Abstract

**Background:** Early detection of suicide risk in adolescents is crucial but faces challenges including stigma, reluctance to disclose suicidal thoughts, and limited accessibility of mental health resources. Traditional assessment methods may miss at-risk populations, particularly in community settings. This study aimed to explore whether multimodal analysis combining acoustic and linguistic features can improve prediction of suicide risk in adolescents.

**Methods:** Voice recordings and transcribed text from 600 Chinese adolescents (aged 10-18 years) were collected from 47 schools in Guangdong, China. Suicide risk labels were derived from the Mini International Neuropsychiatric Interview for Children and Adolescents (MINI-KID). The dataset included three voice tasks: answering an open-ended question about emotional regulation, reading a standard passage, and describing a face with negative emotions. Features were extracted using pre-trained models (EMOTION2VEC for acoustic features, Paraformer for speech-to-text conversion, and Tongyi Qianwen’s text-embedding-v3 for text features). We applied various machine learning classifiers including Support Vector Machine, Multi-layer Perceptron, Random Forest, and XGBoost to develop both single-modal and multimodal prediction models. Front-end fusion (FF) and back-end fusion (BF) techniques were employed to combine acoustic and linguistic features.

**Results:** Fusion models combining both acoustic and linguistic features consistently outperformed individual models. The model with both front-end and back-end fusion achieved the highest performance with an accuracy of 0.73, precision of 0.70, recall of 0.80, and F1 score of 0.74. Front-end fusion alone achieved the highest Area Under the Receiver Operating Characteristic Curve (AUROC) of 0.767. Models performed equivalently across age groups but significantly better in females (AUROC = 0.72) compared to males (AUROC = 0.46).

**Conclusions:** Multimodal analysis combining acoustic and linguistic features significantly improves predictive accuracy for adolescent suicide risk detection compared to single-modal approaches. This approach offers a promising method for early identification of at-risk adolescents in community settings, potentially enabling timely intervention. Further external validation with larger samples is needed to optimize these models for clinical application.

## Background

Suicide is a severe public mental health issue in adolescents. The trend of suicide rate in adolescents by diffrent means increased on average by 1-2.7% annually [1]. The global prevalence of suicidal ideation in the past 12 months was 16.9% in adolescents from low-income and middle-income countries, particularly in the African and the western Pacific regions, and particularly among girls and adolescents aged 15-17 years [2]. Due to the increased suicide rate, there is a pressing need to detect suicidal risks early to prevent severe consequences. Suicidal ideation in adolescents has broad impact on life including increased likelihood of future suicide attempts [3], poor social functioning [4], a worse life and fewer opportunities in adulthood [5]. Timely detection of suicidal risks could help save lives and identify at-risk population for timely intervention.

However, early detection faces a lot of challenges. The sensitive and stigmatized nature of suicide often presents barriers to self-report data collection and clinical interviews. Individuals may be reluctant to share personal or confidential information related to suicide [6]. Obtaining a substantial amount of labeled data about suicidal risks may be limited. Therefore, novel approach is needed to identify suicidal risks at an early stage. Sentiment analysis is a rapidly evolving research field to analyze and understand human emotional states. Traditional sentiment analysis methods mainly rely on written texts. However, as human language is complex, acoustic tone and various voice characteristics may carry richer information and give better insights into the sentiments [7]. Multimodal sentiment analysis that combines data from various modalities such as text and sound can be helpful to identify early signs of suicidal ideation and prevent attempts at suicide.

Recent advances in digital technologies provide the potential to utilize speech information to monitor individuals’ emotional status [8] and psychiatric disorders [9]. However, suicidality is more complex. Some studies have extended the application of multimodal sentiment analysis in the field of suicide research. For example, a study in the US developed a random forest classifier using 588 veterans narrative audio and transcribed text to select 15 acoustic and linguistic features of speech. This model achieved a sensitivity of 0.84, a specificity of 0.70 and an accuracy of 0.72 [10]. Another research used words and vocal characteristics of 379 subjects from clinical settings to develop a support vector machine (SVM)-based classification algorithms with accuracies ranging from 0.74–0.85 [11]. Although these studies have achieved relatively high accuracy, they are based on English language and mainly from the adult population. Few research has focused on the suicide prediction in the adolescents in the community which may include a broader at-risk population according to the iceberg theory of suicidality [12].

In the studies based on Chinese language, a SVM classifier of suicidal risk developed by text analysis using social media demonstrated a sensitivity of 0.64 and a specificity of 0.32 [13]. Although the metrics in people with suicidal-related communication were slightly higher, the results is difficult to be generalized into all respondents. Another study utilized comments in a microblog online group and found that the best overall models were the SVM models with an accuracy of 0.85 [14]. Although this result outperformed other suicide machine learning models to identify posts with suicide content, the sample were mainly single females with a college degree. More evidence is needed in a diverse population of adolescents to identify a high-risk population for lifesaving. Current research in the Chinese language mainly focuses on text features extracted from public posts, other types of data such as speech could also be effective predictors. A recent study used speech audio and transcribed text from Beijing psychological support hotline to develop deep learning models and large language models (LLMs) to predict future suicide behaviour [15]. The combination of LLMs with manual scales achieved an F1 score of 76.47%, a sensitivity of 86.68%, a specificity of 72.73%, and a precision of 68.42%. However, the performance of speech models and the fusion methods still need exploration. Examining the integration of potential factors may provide additional insights to build more effective models to detect suicide risks.

In this study, we analyzed the data from both speech audio and transcribed text using traditional machine learning models and deep learning models. We compared the results from single-modal classifiers and multimodal classifiers using front-end and back-end fusion strategies. To our knowledge, few prior study explored multimodal fusion strategies of speech and text in the integration of deep learning to predict suicidal risk. Our study highlights the way for further exploration of multimodal techniques in suicide prediction and offers promising opportunities for early intervention and support.

## Methods

### Dataset

The data used in this study was collected from 47 elementary and middle schools in Guangdong, China [17]. City-wide screening was conducted to identify at-risk adolescents and non-risk adolescents based on the Mini International Neuropsychiatric Interview for Children and Adolescents (MINI – KID) scale by clinical experts. Invitations for interview were sent through schools and 600 Chinese adolescents aged 10 – 18 were recruited. The interview included three voice tasks: Task 0 required subjects to answer an open - ended question about emotional regulation; Task 1 was to read aloud “The North Wind and the Sun“; and Task 2 was to describe a face with negative emotions using their own words. Interviews were conducted in Mandarin by trained research assistants under the supervision of psychiatrists. Informed consent was obtained from all subjects and their guardians before data collection. All data has been anonymized for the speakers before we accessed, with sensitive information such as names and geographical locations removed. The dataset consists of a training set with 400 samples, a development set with 100 samples and a test set with 100 samples. 50% of the samples having a suicide risk label in both training and development set. As we did not access the labels in test set, only training and development set were used in this study.

### Ethical Consideration

The Data Use Agreement was signed between research team members and Tsinghua University Multimedia Signal and Intelligent Information Processing Lab.

Data was shared solely for research questions of 1st SpeechWellness Challenge Interspeech 2025 [17]. All participants (speakers) had been anonymised before the dataset became available. All analyses conducted in this study aimed to advance scientific knowledge with no commercial or financial conflicts of interest.

### Feature extraction

We extracted feature vectors using pre-trained speech and text models as they are effective to obtain relevant features. We used the fine-tuned version of the pre-trained model EMOTION2VEC [18] on the ModelScope platform to extract speech features, which is the first universal speech emotion representation model. We used the Paraformer-large model [19] on the ModelScope platform to extract the textual content from the audio, and then used Tongyi Qianwen’s text-embedding-v3 model [20] to extract the text features.

### EMOTION2VEC

Emotion2vec is a general - purpose speech emotion representation model based on self - supervised learning [18]. It employs a multi - layer convolutional neural network as a feature extractor and a multi - layer Transformer as the backbone network. We used it to extract features from audio files, obtaining 768 - dimensional feature vectors. We also used its classification model to further obtain 9 - dimensional emotion features. We mainly used the 768 - dimensional feature vectors for binary classification of suicidal risk as it captured a wider range of information in speech.

#### Paraformer

Paraformer is a model for non - autoregressive end - to - end speech recognition [19]. It adopts a joint training method using cross - entropy, mean absolute error, and minimum word error rate to achieve fast and accurate speech recognition. In this study, we used it to transcribe audio to text.

#### Text-embedding-v3

Text - embedding - v3 of Tongyi Qwen is a multilingual unified text vectorization model based on the LLM foundation, launched by Tongyi Lab in July 2024 [20]. In the evaluation of Chinese public retrieval datasets, its retrieval performance is better among similar models. So we chose to input the text obtained from Paraformer into Text - embedding - v3 to get a 1024 - dimensional text feature vector.

Through the above steps, we obtained a 768 - dimensional speech emotion feature vector and a 1024 - dimensional text feature vector respectively. We also had two demographics features: age and gender which are crucial factors associated with suicidal thoughts and behaviors [21]. Furthermore, we also performed direct concatenation and weighted average processing on the speech feature vectors of the three tasks to obtain a 3*768 - dimensional vector and a 768 - dimensional vector respectively, aiming to explore the optimal input.

### Design of single-mode classifiers

We used several different algorithms to perform classification tasks on speech or text features separately. Regarding speech features, we completed the classification tasks for task 0, task 1 and task 2 as well as direct concatenation task and weighted average processing task. As for text features, since the data for task 1 was the same across different scenarios, we completed the classification tasks for task 0 and task 2 respectively.

#### Support Vector Machine (SVM)

In speech feature classification, we found that the SVM method achieved the best prediction results on 9 - dimensional emotion features. Therefore, we combined the 9 - dimensional emotion features with age and gender to form feature vectors. For gender, it is encoded as 0 (male) or 1 (female), while the age uses its original numerical value. Then, we used StandardScaler to adjust the distribution of each feature to a standard normal distribution with a mean of 0 and a standard deviation of 1. The training and development sets were processed based on the statistics of the training set to eliminate the influence of scale differences. For model training, we selected an SVM classifier with a radial basis function (RBF) kernel. By defining a parameter grid containing different values of C ([0.01, 0.1, 1, 10, 100, 1000]), gamma ([0.001, 0.01, 0.1, 1, 10, 100]), and kernel function types. Due to the small number of samples, we used GridSearchCV to conduct a grid search with 5 - fold cross - validation to determine the optimal parameters. In text feature classification, we found that the SVM method yielded poor results, so we did not use it.

#### Multi - layer Perceptron (MLP)

In speech feature classification, we first combined age, gender and speech features, performed 0 - 1 encoding on gender, and then standardized the training and development set data using StandardScaler to eliminate scale differences. The constructed MLP model consisted of an input layer, three hidden layers, and an output layer. The number of neurons in the input layer was determined by the dimension of the input features which was 770 (task 0, task 1, task 2 and weighted average processing task) and 2306 (direct concatenation task). Each layer was equipped with a Rectified Linear Unit (ReLU) activation function to introduce non - linearity, and dropout (with ratios of 0.3, 0.2, and 0.1 respectively) was used to prevent overfitting. During training, the cross - entropy loss function and the Adam optimizer were used. In text feature classification, we performed the same processing on the features. In model construction, the MLP model had an input layer, two hidden layers, and an output layer. The number of neurons in the first and second hidden layers was 1024 and 256 respectively. Each layer was followed by a ReLU activation function and a dropout of 0.4.

#### Random Forest

In speech feature classification, we followed the same feature processing method as that for the MLP. When constructing and tuning the model, we selected the RandomForestClassifier model and set a random seed to ensure the reproducibility of the results. We defined a grid param_grid containing several key hyperparameters for systematic searching. Specifically, n_estimators was set to [100, 200, 300] to explore the influence of different numbers of decision trees on the model, max_depth was set to [None, 10, 20] to control the maximum depth of the decision trees and avoid overfitting or underfitting, and min_samples_split was set to [2, 5, 10] to specify the minimum number of samples required to split an internal node. The remaining parameters were set to their default values. We used GridSearchCV in combination with 5 - fold cross - validation, and searched for hyperparameters with accuracy as the evaluation metric to determine the best model. The same method was used for text feature classification.

#### XGBoost

We replaced the SVM method for text recognition with XGBoost. The feature processing method was the same as that of MLP. The data was converted into the DMatrix format to meet the input requirements of the XGBoost model. The parameters were set as follows: the objective was set to binary:logistic; the eval_metric was chosen as error to use the error rate as the evaluation metric; the eta was set to 0.05 to control the learning step of the model; the max_depth was set to 6 to limit the maximum depth of the decision tree and prevent overfitting; the subsample was set to 0.8 to randomly sample 80% of the samples for training; the colsample_bytree was set to 0.8 to randomly sample 80% of the features for tree construction; the seed was set to 42 to ensure the reproducibility of the results. Meanwhile, an early - stopping mechanism was introduced.

### Front-end and back-end fusion classifier design

#### Front-end fusion(FF)

We adopted the front - end fusion method to integrate speech features and text features and used a MLP model to carry out the classification task. First, we defined the MyDataset class. For each sample, we extracted the age, the gender encoded with 0 - 1, the speech features, and the text feature vector. Since we couldn’t find a good alignment method, these features were directly concatenated into a single feature vector at the front - end to form a fused feature.

Then, we constructed an MLP model consisting of three fully - connected layers. A ReLU activation function is used between each layer to introduce non - linearity, and dropout (with a probability of 0.4) was applied to prevent overfitting. During the training process, the cross - entropy loss function and the Adam optimizer were employed, with a learning rate set to 0.0001. The number of training epochs was 200, and an early - stopping mechanism was adopted with a patience value of 30. The accuracy of the development set was used as the judgment criterion, and the best model was saved. Through front - end fusion, we expected to fully explore the intrinsic relationship between speech features and text features and improve the performance of the classification model with a more comprehensive and rich feature representation. The procedure is outlined in Figure 1.

**Figure 1.**
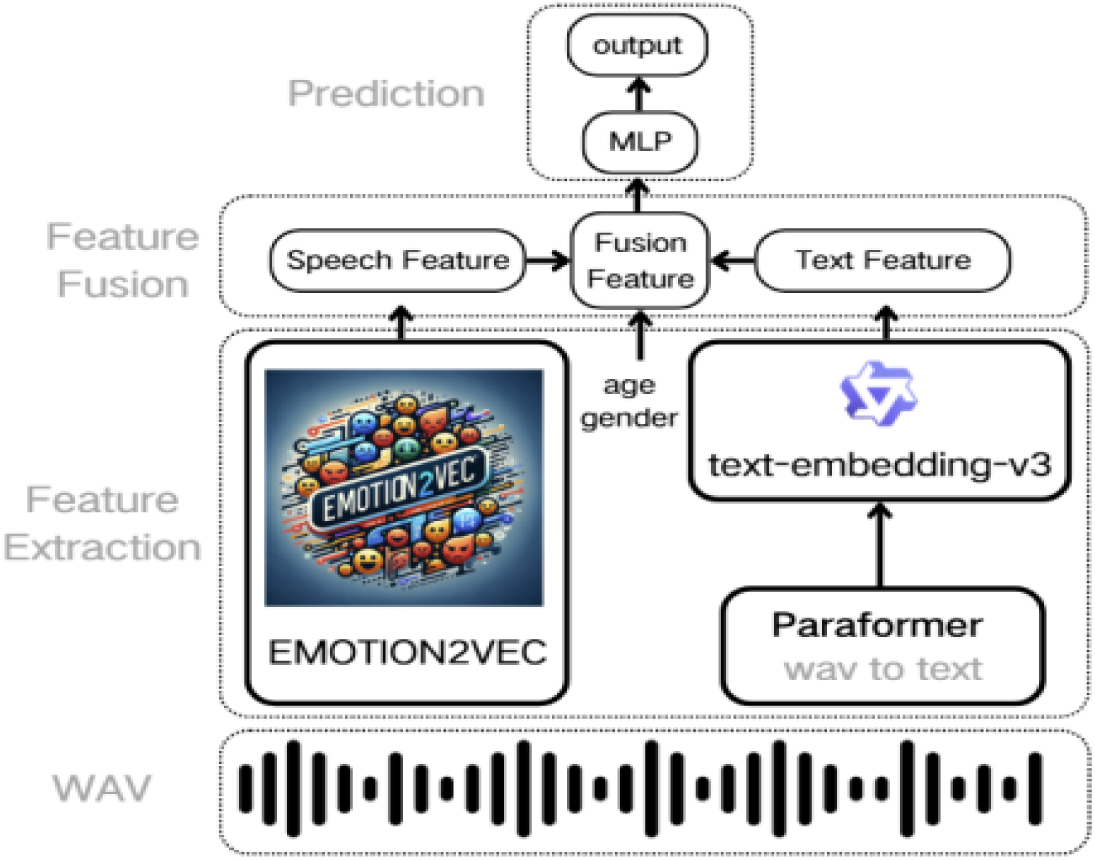
The flowchart of front-end fusion.

#### Back-end fusion(BF)

We also used the back - end fusion approach to integrate the prediction results of each model. We generated all possible weight combinations (with a weight step of 0.02) and determined the optimal weight combination based on the highest average accuracy on the training and development sets. We would like to fully leverage the advantages of different models to improve the overall classification performance. The optimal weight combination we found was as follows: mlp_txt : 0.14 mlp_speech : 0.04 rf_txt : 0.1 svm_speech : 0.72. In addition, we also incorporated the results from 3.4.1(FF) with the purposes to further enhance the classification performance. The optimal weights we found were: FF : 0.4 mlp_txt : 0.14 mlp_speech : 0.04 rf_txt : 0.42. The procedure is outlined in Figure 2.

**Figure 2.**
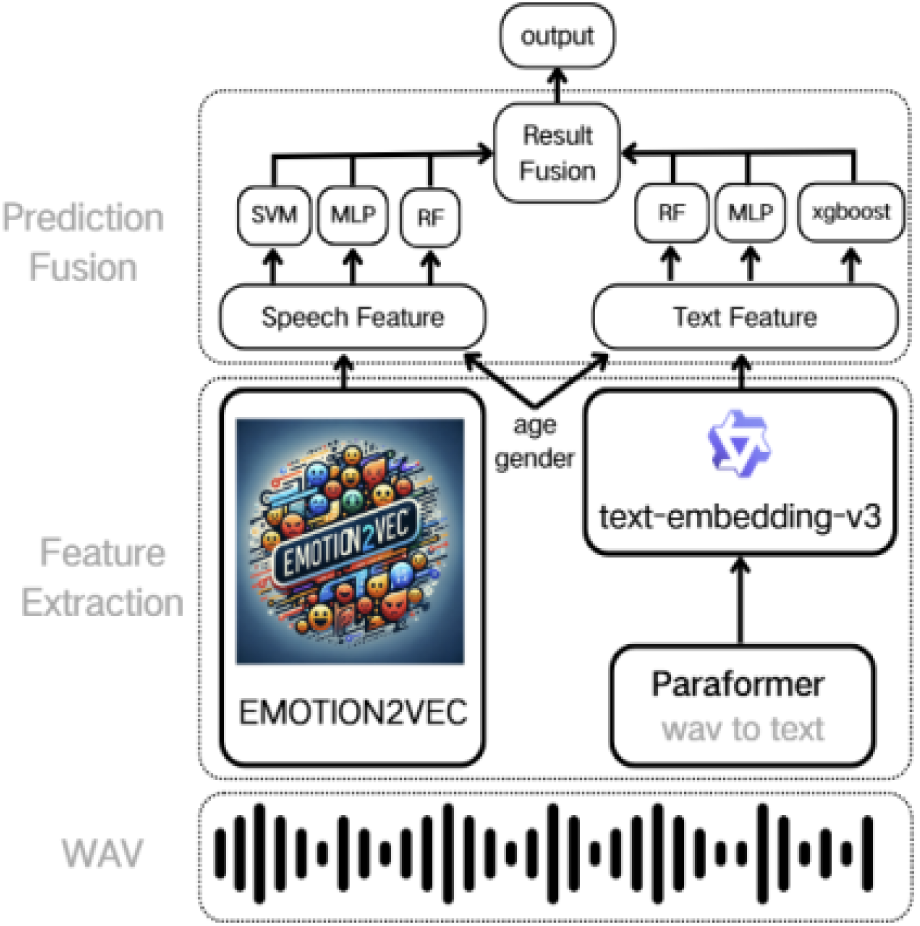
The flowchart of back-end fusion.

## Results

### Demographics and recordings characteristics

The training set consisted of 400 adolescents, with 280 females (70%) and a mean age of 13.77 years (SD = 2.43). In the development set, there were 100 adolescents, with 66 females and 34 males. The mean age of participants in the development set was 13.76 years (SD = 2.44). Scattertext analysis of the natural language in task 0 and task 2 in both training and development set showed the top words used by suicidal and non-suicidal adolescents. We analyzed 44346 words from the original text corpus and also excluded the auxiliary words such as modal verbs to get a text corpus of 30990 words. At-risk adolescents tended to speak more than non-risk counterparts, both before (23,600 vs. 20,746 words) and after (16,411 vs. 14,579 words) exclusion. Top words used by at-risk adolescents include first person singular pronouns, third person singular pronouns, probability, mood, “bad” and “comparison” after excluding the auxiliary words. However, non—risk adolescents showed similar top words. We did not find different patterns of top words in the two groups of adolescents. Detailed information can be seen in Supplementary Figure (a-c) which is available by contacting the corresponding author as they are not in English.

### Prediction performance

The accuracy for each model is presented in Table 1. In acoustic models, task 1 generally achieved the highest accuracy across all models, with scores ranging from 0.45 to 0.62. Specifically, the acoustic MLP model performed best in task 1 with an accuracy of 0.62. However, when combining all tasks, the overall performance did not surpass the accuracy of task 1 alone. This suggests that the inclusion of tasks 0 and 2 did not provide additional benefit and, in fact, led to a slight reduction in accuracy for the acoustic models, as reflected in the combined accuracy scores, which ranged from 0.48 to 0.60. Thus, task 1 appeared to be the most informative feature for predicting the outcome in the acoustic models.

**Table 1.**
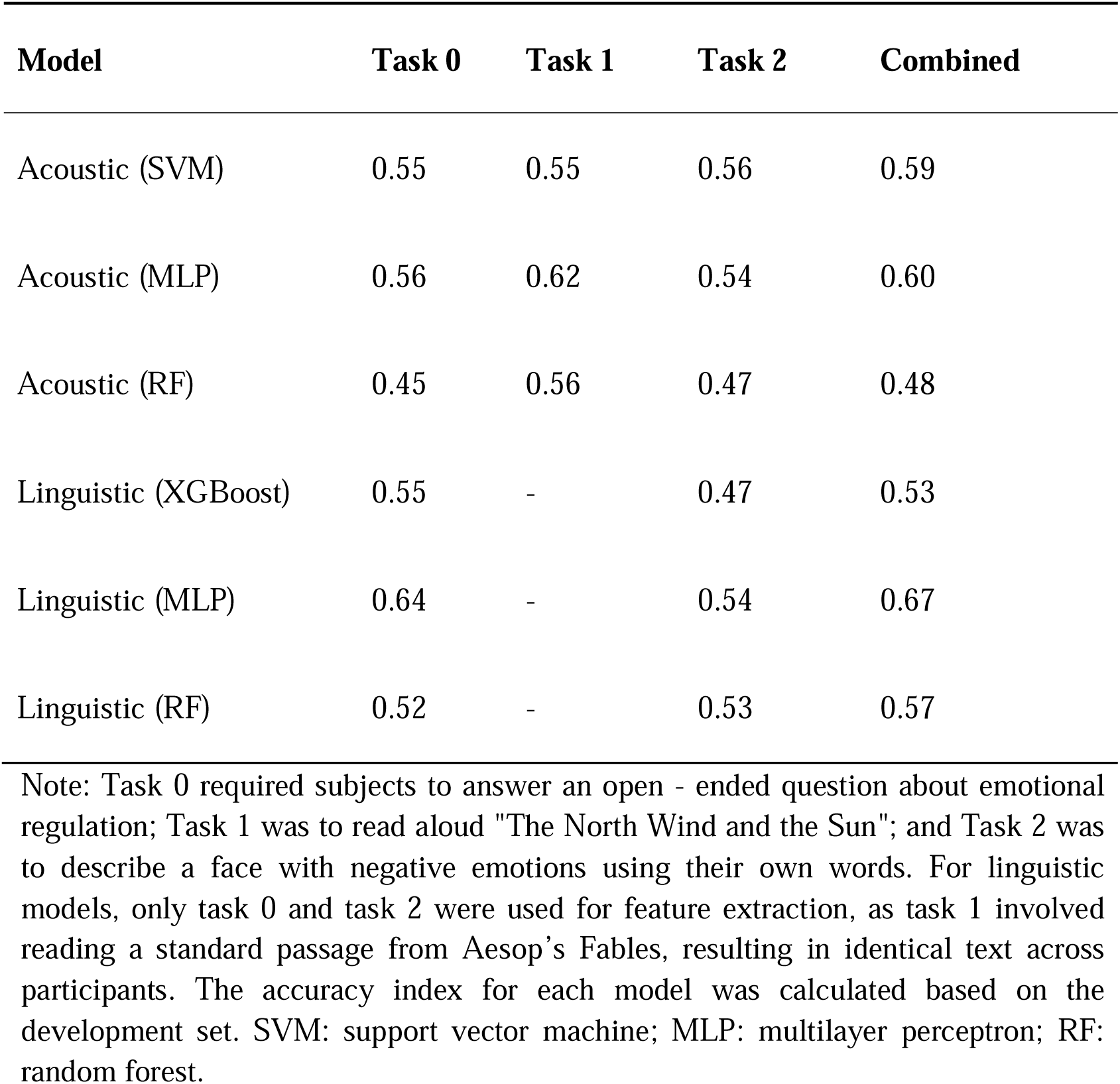
Accuracy for single modal model for each task in the development set.

| Model | Task 0 | Task 1 | Task 2 | Combined |
| --- | --- | --- | --- | --- |
| Acoustic (SVM) | 0.55 | 0.55 | 0.56 | 0.59 |
| Acoustic (MLP) | 0.56 | 0.62 | 0.54 | 0.60 |
| Acoustic (RF) | 0.45 | 0.56 | 0.47 | 0.48 |
| Linguistic (XGBoost) | 0.55 | - | 0.47 | 0.53 |
| Linguistic (MLP) | 0.64 | - | 0.54 | 0.67 |
| Linguistic (RF) | 0.52 | - | 0.53 | 0.57 |

Note: Task 0 required subjects to answer an open - ended question about emotional regulation; Task 1 was to read aloud "The North Wind and the Sun"; and Task 2 was to describe a face with negative emotions using their own words. For linguistic models, only task 0 and task 2 were used for feature extraction, as task 1 involved reading a standard passage from Aesop's Fables, resulting in identical text across participants. The accuracy index for each model was calculated based on the development set. SVM: support vector machine; MLP: multilayer perceptron; RF: random forest.

In the linguistic models, combining tasks 0 and 2 for feature extraction led to better performance than using each task separately. Specifically, when task 0 and task 2 were combined, the linguistic models exhibited improved accuracy compared to using either task individually. For example, the linguistic MLP model achieved a combined accuracy of 0.67, which was higher than the accuracy of 0.64 when only task 0 was used. This suggests that the combination of tasks 0 and 2 provided complementary information that enhanced the model’s ability to predict the outcome. In contrast to the acoustic models, where the combined performance was less effective than task 1 alone, the linguistic models benefited from the synergistic effect of combining task 0 and task 2, underscoring the value of integrating different linguistic tasks for more robust feature representation.

Table 2 presents the prediction performance of models including acoustic, linguistic, and combined acoustic-linguistic models. Among the acoustic models, the MLP model achieved the highest accuracy (0.62), followed by SVM (0.59) and RF (0.56). For linguistic models, the MLP model again outperformed others with an accuracy of 0.67, while the XGBoost model had the lowest accuracy at 0.53. The combined acoustic and linguistic models, including front-end fusion (FF), back-end fusion (BF) and both FF & BF demonstrated improved performance. The FF model achieved an accuracy of 0.70, and BF outperformed FF with an accuracy of 0.71. The best performance was achieved by the FF & BF model, which showed an accuracy of 0.73, precision of 0.70, recall of 0.80, and F1 score of 0.74. Overall, the combined models consistently outperformed the individual acoustic and linguistic models, with FF & BF providing the highest prediction accuracy and F1 score. This suggests that integrating both acoustic and linguistic features through fusion techniques enhances predictive performance across all metrics.

**Table 2.** Prediction performance for single- and multi-modal model in the development set.

|  | Model | Specificity | Sensitivity | Accuracy | Precision | Recall | F1<br>Score |
| --- | --- | --- | --- | --- | --- | --- | --- |
| Acoustic | SVM | 0.54 | 0.66 | 0.59 | 0.61 | 0.59 | 0.57 |
| Acoustic | MLP | 0.48 | 0.76 | 0.62 | 0.59 | 0.76 | 0.66 |
| Acoustic | RF | 0.48 | 0.64 | 0.56 | 0.55 | 0.64 | 0.59 |
| Linguistic | XGBoost | 0.58 | 0.48 | 0.53 | 0.53 | 0.48 | 0.50 |
| Linguistic | MLP | 0.68 | 0.66 | 0.67 | 0.67 | 0.66 | 0.67 |
| Linguistic | RF | 0.64 | 0.50 | 0.57 | 0.58 | 0.50 | 0.54 |
| Acoustic&Linguistic | FF | 0.64 | 0.76 | 0.70 | 0.67 | 0.76 | 0.71 |
| Acoustic&Linguistic | BF | 0.72 | 0.70 | 0.71 | 0.71 | 0.70 | 0.70 |
| Acoustic&Linguistic | FF & BF | 0.66 | 0.80 | 0.73 | 0.70 | 0.80 | 0.74 |

Note: SVM: support vector machine; MLP: multilayer perceptron; RF: random forest; XGBoost: Extreme Gradient Boosting; FF: front-end fusion; BF: back-end fusion.

Figure 3 displays the ROC curves for different models along with their corresponding Area Under the Receiver Operating Characteristic Curve (AUROC) values. The FF model achieved the highest AUROC of 0.767, indicating superior performance, followed closely by FF & BF with an AUROC of 0.723. The BF model had a moderate AUROC of 0.676, while MLP models using text (0.656) and speech (0.550) showed lower AUROC values. The RF model with text features had an AUROC of 0.636, and the RF model with speech features had a relatively lower AUROC of 0.543. The SVM model using speech features showed an AUROC of 0.607, while the XGBoost model with text features had the lowest AUROC of 0.583. This figure highlights that fusion models (FF and FF & BF) generally outperform individual feature models in predicting the outcome. Further subgroup analyses showed that the fusion models achieved relative equivalent performance in both adolescents aged ≥ 14 and < 14 (AUROC = 0.65 vs. 0.70). However, models performed significantly better in females compared to males (AUROC = 0.72 vs. 0.46). The gender disparity in model performance indicated an underrepresentation of males in mental health prediction.

**Figure 3.**
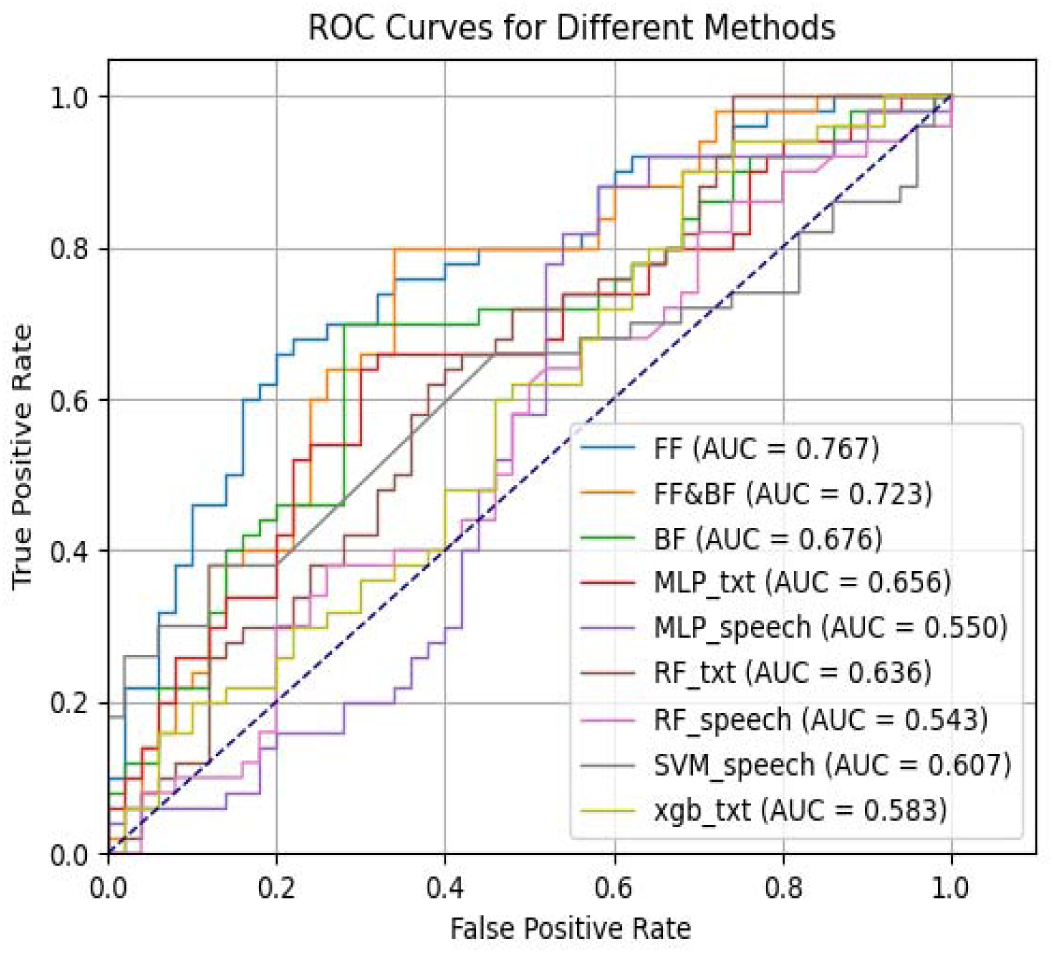
AUROCs for each model.

## Discussion

In this study, we evaluated the performance of single-mode classifiers and front-end and back-end fusion of acoustic and linguistic features in deep learning models to detect suicidal risk in adolescents. The results demonstrate that fusion models, combining both acoustic and linguistic features, consistently outperformed individual models. These findings underscore the importance of incorporating multi-modal features to predict suicidal risk in adolescents.

We did not find significantly different patterns of top words in suicidal and non-suicidal adolescents, although suicidal adolescents tended to speak more than non-suicidal adolescents, which was consistent with previous findings using the text from Chinese social media [13]. This could mainly be explained by the relatively small text corpus in our study. The non-differential word patterns also helped explain the poor performance of models using text features only. Previous research showed various linguistic features as potential predictors of suicidal thoughts and behaviors such as more intensifiers and superlatives and greater usage of pronouns [22]. However, the English-based language may not be directly comparable with Chinese and current evidence is mixed. For example, one study in China used the Simplified Chinese-Linguistic Inquiry and Word Count to group the words into different categories [13]. They did not find a significant association between first person singular pronouns (ie, I, me, and my) and suicide risk, which was inconsistent with earlier research in Chinese context [23]. Although previous scholars has developed a Chinese suicide dictionary [24], it was based on social media setting which was very informative and different from our research context. Therefore, we directly used a unified pre-trained text embedding model to obtain text feature vectors in this study.

As for the classification models of suicidal risk, we found that there were equivalant performances of models based on acoustic features alone and linguistic features alone. Multimodal models with the combination of acoustic, linguistic and demographic features using fusion techniques achieved the best performance. This is not completely in line with previous findings that acoustic-based models performed better (AUROC_=_0.78) than linguistic-based models (AUROC_=_0.74) to detect suicidal ideation in veterans [10]. However, there is consistent evidence in the research of not only suicide but also other mental health status where fusion of different modalities helped improve prediction results [25]. AUROCs reached by our FF and FF & BF models are also comparable to previous research on suicidal risk [10]. Among the very few research using speech data in Chinese, several deep learning approaches such as Recurrent Neural Network, Graph Neural Networks and Mamba were used for the speech data from the psychological support hotline in Beijing [15]. The performance of acoustic models were similar to ours in terms of sensitivity, specificity, precision and F1-score. However, our fusion models achieved much better overall performance. Although they applied LLMs to predict suicidal risk and demonstrated superior performance, another study compared supervised learning and LLMs in the prediction of suicidal risks in Chinese social media and found that LLMs may fail in some tasks leading to overprediction and misclassification [26]. Caution is needed to adopt innovative artificial intelligence approaches in mental health domains.

This study has several limitations. First, the simplification of suicidal risk to binary labels does not consider that suicidality might exist on a spectrum [27]. The classification reflected participants’ responses to the MINI-KID scale which assesses current suicide risk and should not be interpreted as a prediction of future suicidal behavior. Second, only the speech and text of tasks as well as age and gender are available in this study. There could be other omitted variables which are important to predict suicidal risks in adolescents. Third, this study was conducted in a relatively small sample size, evidence is needed to expand the development and test set to achieve more reliable results.

## Conclusions

This study showed that multi-modal machine learning models is a promising approach for detecting suicidal risk in adolescents. Models using combined acoustic and linguistic features consistently performed better than single-modal models. Further external validation and optimization are needed to validate and improve the results on unseen data.

## Data Availability

All data produced in the present study are not available based on the Data Use Agreement between research team members and Tsinghua University Multimedia Signal and Intelligent Information Processing Lab.

## List of abbreviations

AUROC: Area Under the Receiver Operating Curve
BF: Back-end Fusion
FF: Front-end Fusion
LLM: Large Language Model
MINI-KID: Mini International Neuropsychiatric Interview for Children and Adolescents
MLP: Multi - layer Perceptron
ReLU: Rectified Linear Unit
RF: Random Forest
SVM: Support Vector Machine
XGBoost: Extreme Grading Boost

